# Diagnostic accuracy of automated Lumipulse plasma pTau-217 in Alzheimer’s disease

**DOI:** 10.1101/2024.05.20.24307613

**Authors:** Giordano Cecchetti, Federica Agosta, Giulia Rugarli, Edoardo Gioele Spinelli, Alma Ghirelli, Matteo Zavarella, Ilaria Bottale, Francesca Orlandi, Roberto Santangelo, Francesca Caso, Giuseppe Magnani, Massimo Filippi

## Abstract

**INTRODUCTION:** Considerable advancements have occurred in blood-based Alzheimer’s disease (AD) biomarkers, with automated assays emerging for clinical use. Demonstrating the reliability of these automated systems is crucial with upcoming AD therapies.

**METHODS:** This cross-sectional study in a Memory Center enrolled 98 patients along the AD continuum or affected by other neurodegenerative disorders, stratified by CSF A/T status and clinical syndrome. Plasma pTau-217, pTau-181, and Aβ42/Aβ40 were measured using Lumipulse. Relationships with CSF and glomerular filtration rate (GFR) were explored. ROC analysis was conducted to assess diagnostic performance.

**RESULTS:** GFR effect was lowered by the use of ratios and pTau-217 correlation with CSF was strong. Plasma pTau-217 discriminated A+/T+ status with excellent accuracy in both dementia and mild cognitive impairment (AUC 0.93-0.97), outperforming pTau-181 and Aβ42/Aβ40. Cutoffs displayed high diagnostic performance.

**DISCUSSION:** Lumipulse automated pTau-217 and identified cutoffs exhibit excellent diagnostic accuracy for CSF A+/T+ status and clinical group detection, facilitating future clinical translation.

## 1. Background

The urgent need for disease-modifying therapies for Alzheimer’s disease (AD) underscores the importance of accurate biological diagnosis. As per the National Institute on Aging and Alzheimer’s Association (NIA-AA) criteria established in 2018,^1^ AD can be diagnosed biologically, regardless of the clinical presentation, through the detection of biomarkers indicative of both amyloidopathy (A: CSF Aβ42/Aβ40 and/or amyloid PET) and tauopathy (T: CSF pTau-181 and/or Tau PET). In the last decade, significant attention has been directed towards analyzing specific plasma biomarkers for AD, including pTau-217, pTau-181, pTau-231, and the Aβ42/Aβ40 ratio. These biomarkers have demonstrated considerable correlations with amyloid and tau pathology across the AD continuum, exhibiting considerable accuracy in distinguishing AD from other clinical conditions.^2–4^

Various assays have been developed for measuring AD blood-based biomarkers,^5^ with a recent focus on automated platforms due to their broader diagnostic coverage, ease of use, and cost-effectiveness compared to ultra-sensitive assays.^6, 7^ Among these options, the fully automated Lumipulse G600II assays for pTau-181 and Aβ42/Aβ40 ratio have recently been described in a large sample of both cognitively impaired and unimpaired patients, with pTau-181 showing the highest accuracy in differentiating A+/T+ subjects from other CSF classes (AUC: 0.88-0.91).^3^ Notably, a number of studies have consistently highlighted the higher discriminative performance of plasma pTau-217 over other pTau isoforms,^7–9^ leading the Alzheimer’s Association workgroup to potentially include it among core biomarkers for AD diagnosis.^10^

In light of the recent availability of Lumipulse kits for the measurement of plasma pTau-217, this study aims to assess the discriminative performances of plasma pTau-217 and pTau-217/Aβ42 ratio compared with pTau-181 and Aβ42/Aβ40 ratio across CSF A/T classes and diagnostic groups within a memory-center-based population of cognitively impaired patients. Such investigation is crucial for advancing our understanding of the diagnostic utility of these blood-based biomarkers in the context of AD diagnosis and may have implications for future clinical practice.

## 2. Methods

### 2.1 Participants

The study prospectively recruited patients with cognitive disturbances who underwent diagnostic CSF lumbar puncture (LP) for AD biomarkers measurement at the Neurology Unit of IRCCS San Raffaele Scientific Institute between October 2023 and May 2024. The inclusion criteria comprised: i) a clinical diagnosis of mild cognitive impairment (MCI) or dementia at discharge, ii) detailed clinical/neurological evaluation and neuropsychological assessment within 6 months from LP, iii) complete blood tests, iv) brain imaging scan (either CT or MRI), and v) Apolipoprotein E genotyping. Selected patients also underwent ^18^F-FDG PET and/or DaT-Scan, based on clinical judgment. Glomerular filtration rate (GFR) were calculated for all patients and kidney disease (KD) was defined as having a GFR < 60 ml/min/1.73 m^2^.^3^

Based on CSF Aβ42/40 and pTau-181 patients were stratified according to their A/T profile.^1, 3^

Patients were further categorized into four diagnostic classes based on their clinical diagnosis at hospital discharge: dementia due to AD (AD-DEM), mild cognitive impairment due to AD (AD-MCI), mild cognitive impairment not related to AD (NonAD-MCI), and dementia not related to AD (NonAD-DEM, including Lewy body dementia, normal pressure hydrocephalus, vascular dementia, behavioral variant of frontotemporal dementia, progressive supranuclear palsy, and Parkinson’s disease dementia). This classification facilitated a clearer demonstration of the discriminative accuracy of plasma biomarkers between the AD and other neurodegenerative diseases while ensuring adequate sample sizes.

The ethical standards committee on human experimentation of IRCCS San Raffaele Scientific Institute (Milan, Italy) approved the study protocol and all participants provided written informed consent prior to study inclusion. The study was performed in accordance with the ethical standards as laid down in the 1964 Declaration of Helsinki and its later amendments.

### 2.2 CSF and plasma collection and biomarkers measurement

CSF samples were collected in the morning in sterile polypropylene tubes and levels of CSF Aβ42, Aβ40 and pTau-181 were calculated according to standardized procedures.^11^

Blood withdrawal for plasma collection was performed in dipotassium EDTA anticoagulant tubes through venipuncture right before LP. Blood samples were then centrifugated for 10 minutes at 2000 g, plasma aliquoted in 1 mL portions in polypropylene tubes and stored at –80◦C until use. After being thawed at room temperature for 30 minutes, plasma samples were vortexed for 10 seconds and centrifugated for 5 minutes at 2000 g just before the analysis. Levels of plasma Aβ42, Aβ40, pTau-181 and pTau-217 were measured using fully automated CLEIA on the LUMIPULSE G600II system according to the manufacturer guidelines, as previously described.^3^

### 2.3 Statistical analysis

Analyses were computed using R software with the level of significance set at p < 0.05. Age and CSF biomarkers were compared among CSF A/T and clinical groups using Dunn’s test with Bonferroni correction for multiple comparisons. Fisher’s exact test explored possible differences in prevalence of sex, KD and ApoE ε4 status.

Spearman’s correlation assessed the relationship of plasma biomarkers (pTau-217, pTau-181, Aβ42/Aβ40 ratio, pTau-217/Aβ42 ratio) with CSF biomarkers, age and GFR. Correlation analyses were performed both in the whole sample and CSF A+/A- subgroups, with Bonferroni correction applied for multiple comparisons.

Logistic regression was conducted on rank-transformed plasma biomarkers to assess differences among CSF A/T groups and clinical groups, considering age, sex, and KD as covariates, with Bonferroni correction for multiple comparisons. Subsequently, ROC analysis evaluated the diagnostic performance of non-transformed plasma biomarker values in distinguishing CSF A/T groups and clinical groups, with bootstrap method (n replicates = 2.000) used to assess confidence intervals for the area under the ROC curve (AUC). Bootstrap method (n replicates = 10.000) was also applied to explore differences in the (partial) AUC of the paired ROC curves. Optimal cutoffs of plasma pTau-217 and pTau-217/Aβ42 in differentiating CSF A/T groups were calculated considering the maximum sum of sensitivity and specificity as the metric.

## 3. Results

### 3.1 Sample characteristics

We enrolled a total of 98 patients. Among the CSF A/T profiles, 49 were categorized as A+/T+, 8 as A+/T–, and 41 as A–/T–, with none exhibiting an A-/T+ profile. Across clinical syndromes, there were 27 cases of AD-DEM, 23 of AD-MCI, 15 of NonAD-DEM, and 33 of NonAD-MCI. Demographic and clinical features of different A/T and clinical groups are summarized in Table 1. The patient groups exhibited similar distributions in terms of age, sex, and prevalence of KD. However, ApoE ε4 status showed non-uniform distribution across study groups, being more frequent in the A+ and A+/T+ groups, as well as in clinical groups within the AD continuum.

**Table 1.** Demographic, clinical and biomarkers data of patients stratified according to CSF A and A/T status, and to the clinical syndrome.

|  | N | Age (y) | Sex<br>[F \| M,<br>% F] | KD<br>[+ \| -,<br>%] | ApoE ε4<br>[+ \| -,<br>%] | MMSE | CSF<br>Aβ42/ Aβ 40<br>ratio | CSF<br>pTau-181 | Plasma<br>pTau-217<br>[pg/mL] | Plasma<br>pTau-181<br>[pg/mL] | Plasma<br>Aβ42<br>[pg/mL] | Plasma<br>Aβ40<br>[pg/mL] | Plasma<br>pTau-217/<br>Aβ42 ratio | Plasma<br>Aβ42/Aβ40<br>ratio |
| --- | --- | --- | --- | --- | --- | --- | --- | --- | --- | --- | --- | --- | --- | --- |
| A- | 41 | 70.01 ± 8.15<br>(51.54–83.56) | 20 \| 21<br>(48.8) | 12 \| 29<br>(29.3 %) | 8 \| 33<br>(19.5)* | 24.68 ± 4.84<br>(11.70–30.0) | 0.088 ± 0.009<br>(0.069–0.106) | 28.6 ± 10.3<br>(11.3–51.8) | 0.12 ± 0.06<br>(0.05–0.33) | 1.50 ± 0.65<br>(0.76–3.17) | 28.55 ± 7.31<br>(8.87–50.22) | 324.0 ± 97.4<br>(117.7–700.8) | 0.004 ± 0.003<br>(0.002–0.015) | 0.090 ± 0.015<br>(0.049–0.122) |
| A+ | 57 | 71.68 ± 7.07<br>(53.26–84.04) | 26 \| 31<br>(45.6) | 18 \| 39<br>(31.6 %) | 34 \| 23<br>(59.6)* | **22.73 ± 3.83**<br>**(13.30–29.00)**<br>† | **0.044 ± 0.010**<br>**(0.025–0.066)**<br>† | **102.7 ± 50.3**<br>**(24.3–283.6)**<br>† | **0.70 ± 0.50**<br>**(0.09–2.16)**<br>† | **2.54 ± 1.18**<br>**(0.94–6.25)**<br>† | **24.64 ± 5.18**<br>**(10.86–42.93)**<br>† | 314.5 ± 62.9<br>(142.0–499.6) | **0.031 ± 0.027**<br>**(0.004–0.148)**<br>† | **0.079 ± 0.010**<br>**(0.055–0.099)**<br>† |
| A-/T- | 41 | 70.01 ± 8.15<br>(51.54–83.56) | 20 \| 21<br>(48.8) | 12 \| 29<br>(29.3 %) | 8 \| 33<br>(19.5)* | 24.68 ± 4.84<br>(11.70–30.0) | 0.088 ± 0.009<br>(0.069–0.106) | 28.6 ± 10.3<br>(11.3–51.8) | 0.12 ± 0.06<br>(0.05–0.33) | 1.50 ± 0.65<br>(0.76–3.17) | 28.55 ± 7.31<br>(8.87–50.22) | 324.0 ± 97.4<br>(117.7–700.8) | 0.004 ± 0.003<br>(0.002–0.015) | 0.090 ± 0.015<br>(0.049–0.122) |
| A+/T- | 8 | 71.82 ± 8.15<br>(57.39–83.78) | 4 \| 4<br>(50.0) | 3 \| 5<br>(37.5) | 4 \| 4<br>(50.0)* | 24.70 ± 2.76<br>(21.00–28.00) | **0.051 ± 0.007**<br>**(0.044–0.066)**<br>‡ | 42.9 ± 11.8<br>(24.3–55.4) | **0.33 ± 0.28**<br>**(0.09–0.96)**<br>‡ | 2.35 ± 1.74<br>(1.16–6.25) | **23.49 ± 3.45**<br>**(19.37–28.11)**<br>‡ | 319.7 ± 72.5<br>(250.2–465.8) | **0.014 ± 0.012**<br>**(0.005–0.043)**<br>‡ | **0.075 ± 0.011**<br>**(0.060–0.096)**<br>‡ |
| A+/T+ | 49 | 71.66 ± 6.98<br>(53.26–84.04) | 22 \| 27<br>(44.9) | 15 \| 34<br>(30.6) | 30 \| 19<br>(61.2)* | **22.41 ± 3.90**<br>**(13.30–29.00)**<br>‡ | **0.043 ± 0.010**<br>**(0.025–0.064)**<br>‡ | **112.5 ± 47.3**<br>**(59.4–283.6)**<br>‡ § | **0.75 ± 0.50**<br>**(0.11–2.16)**<br>‡ | **2.58 ± 1.08**<br>**(0.94–5.33)**<br>‡ | **24.82 ± 5.61**<br>**(10.86–42.93)**<br>‡ | 313.6 ± 62.0<br>(142.0–499.6) | **0.033 ± 0.028**<br>**(0.004–0.148)**<br>‡ | **0.079 ± 0.010**<br>**(0.055–0.099)**<br>‡ |
| AD-DEM | 27 | 70.90 ± 7.32<br>(53.30–83.78) | 12 \| 15<br>(44.4) | 11 \| 16<br>(40.7) | 15 \| 12<br>(55.6)* | **19.74 ± 2.90**<br>**(13.30–24.30)**<br>¶ ** | **0.042 ± 0.010**<br>**(0.025–0.057)**<br>¶ †† | **124.0 ± 57.9**<br>**(48.6–283.6)**<br>¶ †† | **0.92 ± 0.55**<br>**(0.23–2.16)**<br>¶ †† | **2.95 ± 1.18**<br>**(1.37–5.33)**<br>¶ †† | 24.07 ± 5.28<br>(10.86–35.03) | 313.5 ± 68.6<br>(142.0–465.8) | **0.042 ± 0.032**<br>**(0.008–0.148)**<br>¶ †† | **0.078 ± 0.012**<br>**(0.055–0.099)**<br>¶ |
| AD-MCI | 23 | 73.07 ± 6.84<br>(53.26–84.04) | 10 \| 13<br>(43.5) | 5 \| 18<br>(21.7) | 15 \| 8<br>(65.2)* | 25.47 ± 2.19<br>(22.00–29.00) | **0.044 ± 0.009**<br>**(0.032–0.064)**<br>¶ †† | **96.3 ± 26.0**<br>**(59.8–145.7)**<br>¶ †† | **0.53 ± 0.32**<br>**(0.11–1.21)**<br>¶ †† | **2.17 ± 0.77**<br>**(0.94–4.58)**<br>¶ | 25.85 ± 5.47<br>(12.80–42.93) | 320.4 ± 61.9<br>(175.2–499.6) | **0.022 ± 0.016**<br>**(0.004–0.078)**<br>¶ †† | 0.081 ± 0.008<br>(0.064–0.095) |
| NonAD-DEM | 15 | 71.17 ± 7.33<br>(56.34–80.73) | 9 \| 6<br>(60.0) | 4 \| 11<br>(26.6) | 3 \| 12<br>(20.0)* | **19.61 ± 4.73**<br>**(11.70–26.00)**<br>¶ ** | 0.082 ± 0.013<br>(0.050–0.101) | 29.0 ± 13.4<br>(11.3–55.4) | 0.16 ± 0.09<br>(0.05–0.33) | 2.04 ± 1.36<br>(0.88–6.25) | 28.05 ± 5.94<br>(18.46–41.79) | 322.1 ± 61.0<br>(226.7–429.9) | 0.006 ± 0.003<br>(0.002–0.013) | 0.088 ± 0.014<br>(0.066–0.110) |
| NonAD-MCI | 33 | 69.51 ± 8.24<br>(51.54–83.56) | 15 \| 18<br>(45.5) | 10 \| 23<br>(30.3) | 9 \| 24<br>(27.3)* | 27.10 ± 1.64<br>(23.40–30.00) | 0.083 ± 0.017<br>(0.044–0.106) | 31.3 ± 10.8<br>(11.6–54.1) | 0.14 ± 0.16<br>(0.06–0.96) | 1.40 ± 0.56<br>(0.76–3.00) | 27.57 ± 7.70<br>(8.87–50.22) | 319.5 ± 103.6<br>(117.7–700.8) | 0.006 ± 0.007<br>(0.002–0.042) | 0.088 ± 0.016<br>(0.049–0.122) |

Values are frequencies (%) or means±standard deviations (min/max). The level of significance was set at $p < 0.05$ and refers to Dunn's test, followed by post-hoc pairwise comparisons (Bonferroni-corrected for multiple comparisons), or to age-, sex- and KD-adjusted linear regression models, or to Chi-squared test (see the main text for further details). Sex and KD were equally distributed among study groups, whereas ApoE ε4 status was differently distributed (\*). Significant comparisons of continuous variables are highlighted in bold and refer to: † = vs A-; ‡ = vs A-/T-; § = vs A+/T-; ¶ = vs NonAD-MCI; \*\* = vs AD-MCI; †† = vs NonAD-DEM. Abbreviations: AD-DEM = dementia due to Alzheimer's disease, AD-MCI = MCI due to Alzheimer's disease, ApoE = Apolipoprotein E gene, KD = kidney disease, NonAD-DEM = dementia non-AD related, NonAD-MCI= MCI non-AD related.

### 3.2 Plasma biomarkers correlation with CSF biomarkers, age and GFR

Spearman’s coefficients and p-values assessing correlations between plasma biomarkers, age, CSF data, and GFR in the entire sample and within A+/A- groups are detailed in Figure 1.

**Figure 1.**
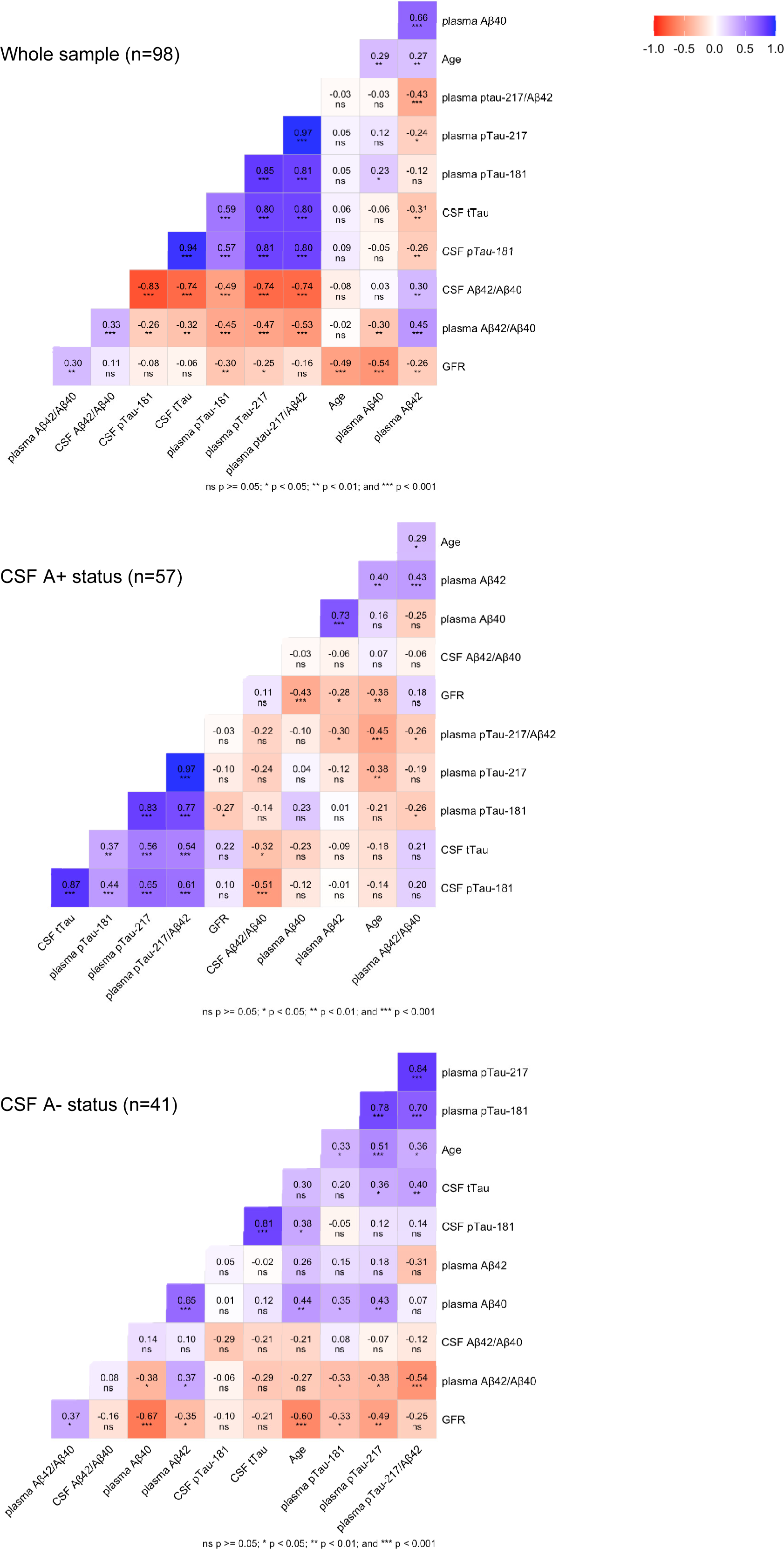
Correlation matrix based on Spearman’s test between plasma biomarkers and age, CSF biomarkers and glomerular filtration rate (GFR). Values represent Spearman’s correlation coefficients (R) and corresponding p-values for the entire sample and for subgroups of patients categorized by CSF A status. The level of significance was set at p < 0.05 and analyses were Bonferroni-corrected for multiple comparisons.

In the whole sample, weak or nonsignificant correlations were observed between plasma biomarkers and age. However, Aβ40 exhibited a moderate negative correlation with GFR (R=-0.54; p<0.001). Notably, plasma pTau-217 and pTau-217/Aβ42 ratio showed strong to very strong correlations with CSF pTau-181 (R=0.81 and R=0.80, respectively; p< 0.001), CSF total Tau ( R=0.80; p<0.001), and CSF Aβ42/Aβ40 (R=-0.074; p<0.001). Plasma pTau-181 displayed similar moderate correlations (∼ CSF pTau-181: R=0.57; ∼ CSF tTau: R=0.59; ∼ CSF Aβ42/Aβ40: R=-0.45; p<0.001). Other explored correlations were nonsignificant or weak.

Within the A+ group, plasma pTau-217 and pTau-217/Aβ42 ratio demonstrated moderate to strong correlations with CSF pTau-181 (R=0.65 and R=0.61, respectively; p<0.001) and total Tau (R=0.56 and R=0.54, respectively; p<0.001). Plasma pTau-181 showed a moderate positive correlation with CSF pTau-181 only (R=0.44; p<0.001). Additionally, plasma pTau-217/Aβ42 ratio and Aβ42 moderately correlated with age (R=-0.45 and R=0.40, respectively; p<0.01). Consistent with the whole group analysis, Aβ40 exhibited a moderate negative correlation with GFR (R=-0.42; p<0.001). Other explored correlations were nonsignificant or weak.

Within the A- group, apart from a moderate positive correlation between plasma pTau-217/Aβ42 and CSF total Tau (R=0.040; p=0.01), nonsignificant or weak correlations emerged between plasma and CSF biomarkers. Plasma pTau-217 and Aβ40 correlated with age (R=0.51 and R=0.44, respectively; p<0.01) and GFR (R=-0.49 and R=-0.67; p<0.01), with moderate strength. Other explored correlations were nonsignificant or weak.

### 3.3 Plasma biomarkers across A/T groups

Figure 2 presents boxplots illustrating the plasma levels of pTau-217, pTau-181, Aβ42/Aβ40 and pTau-217/Aβ42 ratios across CSF A/T groups of patients, alongside with p-values adjusted for age, sex, and KD. Significant differences were observed between A+/T+ and A-/T- subjects, with A+/T+ patients exhibiting higher levels of plasma pTau-217, pTau-181, and p-Tau217/Aβ42, along with lower Aβ42/Aβ40 values. Similar trends were noted in comparisons between A+ and A- patients. Additionally, A+/T- subjects displayed significantly lower levels of pTau-217 and pTau-217/Aβ42 compared to A+/T+ and higher levels compared to A-/T- patients; A+/T- also exhibited lower Aβ42/Aβ40 values than A-/T-. Other comparisons yielded nonsignificant results.

**Figure 2.**
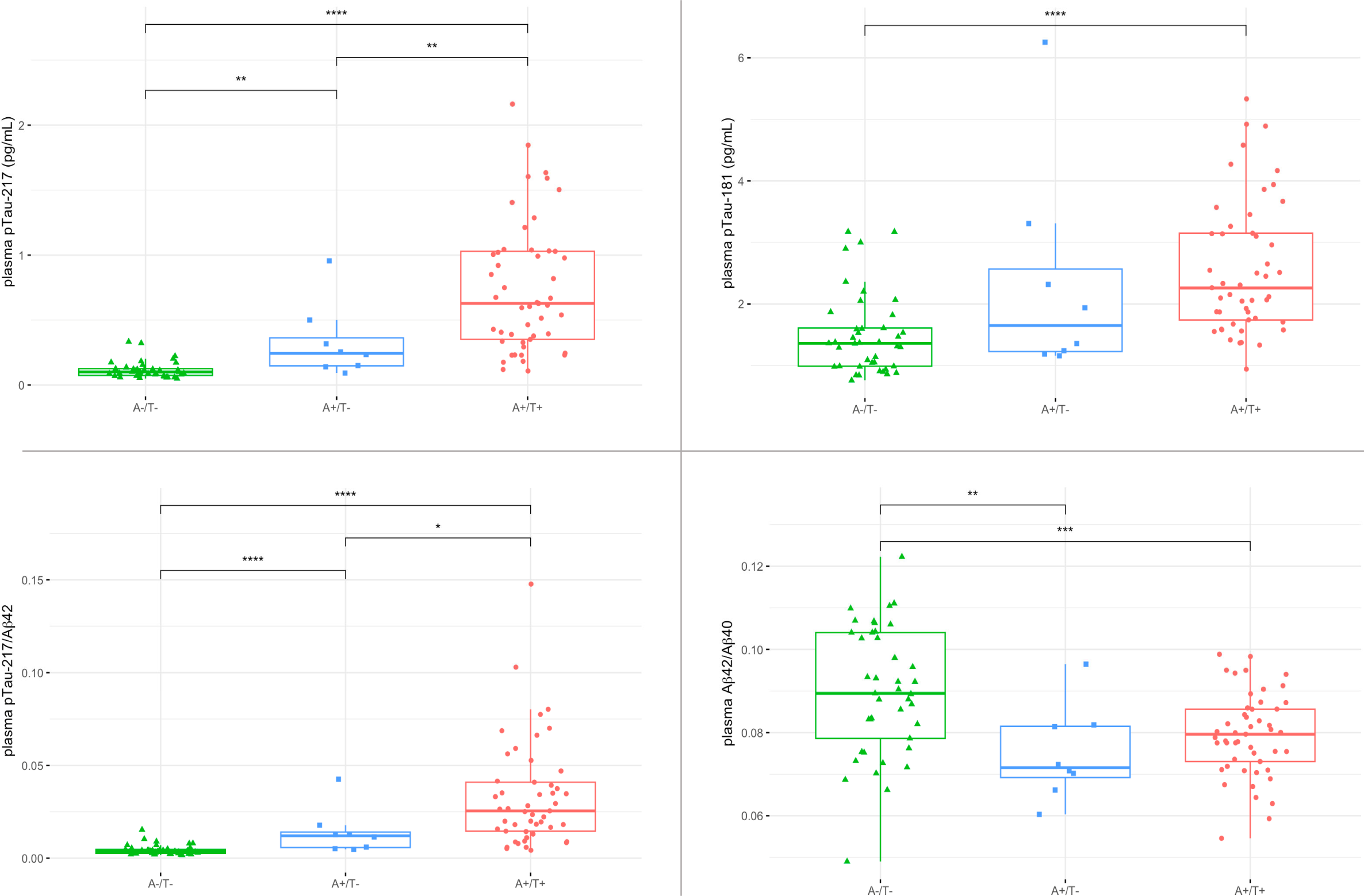
Plasma concentrations of Alzheimer’s disease (AD) biomarkers (pTau-217, pTau-181, pTau-217/Aβ42 ratio, Aβ42/Aβ40 ratio) across the three CSF A/T categories: A-/T-, A+/T-, and A+/T+. The boxplots illustrate the data, with boxes representing the interquartile range, median concentrations depicted by horizontal lines within the boxes, and whiskers extending to the first/third quartile +/- 1.5 times the interquartile range. Pairwise comparisons were conducted on rank-transformed values using logistic regression, adjusting for age, sex, and kidney disease, with p-values Bonferroni-corrected for multiple comparisons.

ROC analysis confirmed the abovementioned differences in plasma biomarkers (Figure 3). Plasma pTau-217/Aβ42 and pTau-217 showed the best performances in all explored comparisons, showing excellent AUC values. Specifically, AUC values ranged from 0.94 to 0.97 in the ‘A+/T+ *vs* A-/T-‘, ‘A+/T+ *vs* not A+/T+’ and ‘A+ vs A-‘ comparisons, and from 0.87 to 0.91 in the ‘A+/T- *vs* A-/T-‘ comparison. The AUCs of plasma pTau-217 and pTau-217/Aβ42 were comparable, and both were significantly greater than those of plasma pTau-181 (AUCs: 0.67-0.83) and Aβ42/40 (AUCs: 0.66-0.81) in all comparisons except for ‘A+/T- *vs* A-/T-’, where Aβ42/40 showed comparable AUC values to those of pTau-217 and pTau-217/Aβ42. Additionally, the AUC of plasma pTau-181 exceeded that of Aβ42/Aβ40 in the ‘A+/T+ vs A-/T-’ and ‘A+/T+ *vs* non-A+/T+’ comparisons. Including KD, sex, age and ApoE status in the linear models did not increase the AUC values of the biomarkers explored (data not shown).

**Figure 3.**
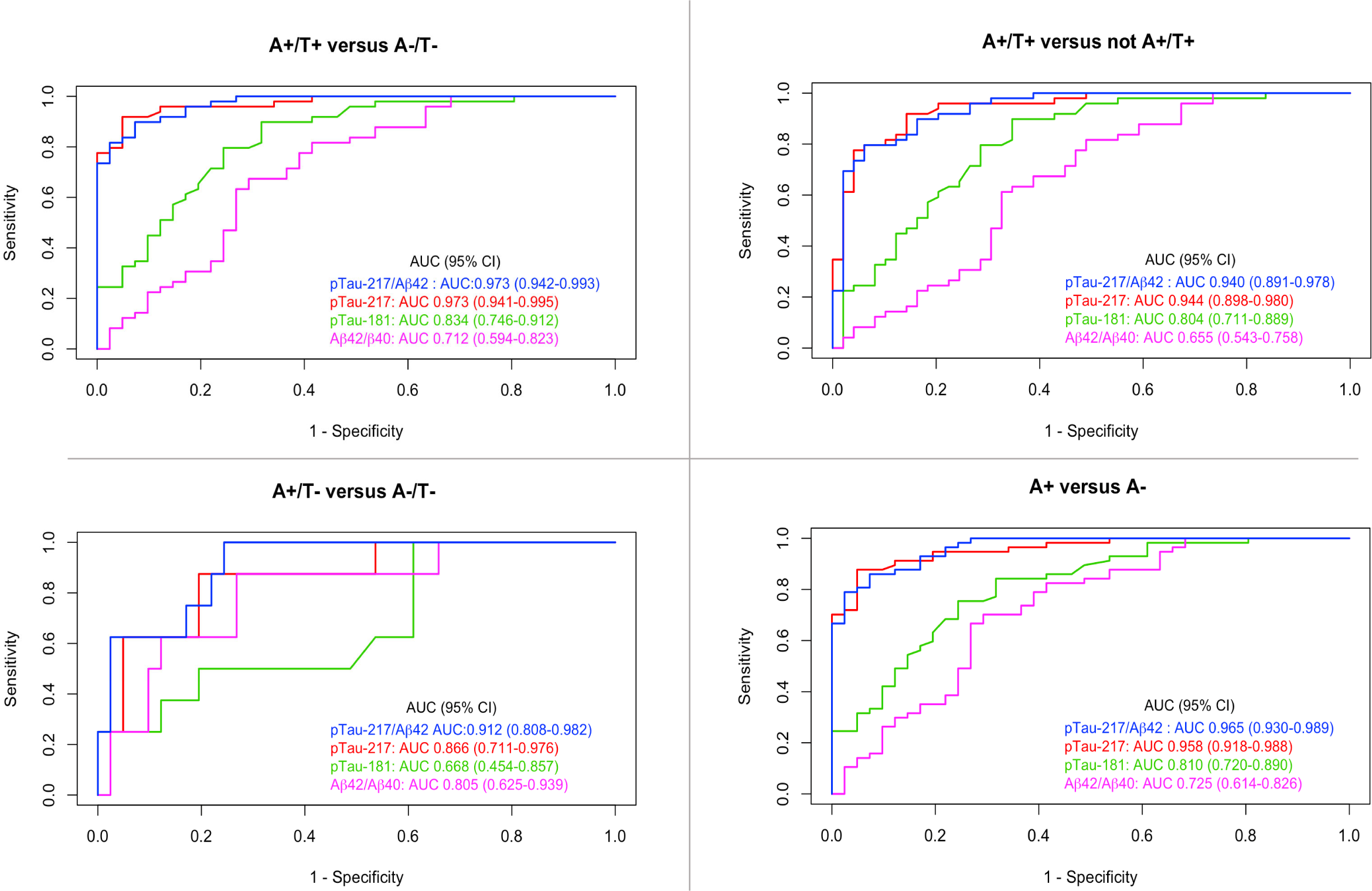
Receiver operating characteristic (ROC) curves assessing the discriminative ability of plasma biomarkers (pTau-217, pTau-181, pTau-217/Aβ42 ratio, Aβ42/40 ratio) in distinguishing among CSF A/T profiles. Areas under the ROC curves (AUC) were calculated for each comparison, with 95% confidence intervals generated by 2,000 bootstrap replicates. Plasma pTau-217 and the pTau-217/Aβ42 ratio demonstrated excellent and superior diagnostic performance in all comparisons compared to pTau-181 and Aβ42/Aβ40.

Optimal cutpoints for plasma pTau-217 and pTau-217/Aβ42, accuracy, sensitivity, specificity, PPV and NPV in explored pairwise comparisons are reported in Table 2.

**Table 2.** Optimal cutoffs of pTau-217 and pTau-217/ Aβ42 in differentiating CSF A/T profiles and their accuracy, sensitivity, specificity, positive and negative predictive values.

|  |  | **Cutoff**<br>**[pg/mL]** | **Accuracy**<br>**[%]** | **Sensitivity**<br>**[%]** | **Specificity**<br>**[%]** | **PPV**<br>**[%]** | **NPV**<br>**[%]** |
| --- | --- | --- | --- | --- | --- | --- | --- |
| **A+/T+**<br>**vs**<br>**A-/T-** | pTau-217 | 0.23 | 93.3 | 91.8 | 95.1 | 95.7 | 90.7 |
| | pTau-217/<br>Aβ42 | 0.0084 | 91.1 | 89.8 | 92.7 | 93.6 | 88.4 |
| **A+/T+**<br>**vs**<br>**not A+/T+** | pTau-217 | 0.23 | 88.8 | 91.8 | 85.7 | 86.5 | 91.3 |
| | pTau-217/<br>Aβ42 | 0.0107 | 84.7 | 81.6 | 87.8 | 87.0 | 82.7 |
| **A+**<br>**vs**<br>**A-** | pTau-217 | 0.23 | 90.8 | 87.7 | 95.1 | 96.2 | 81.6 |
| | pTau-217/<br>Aβ42 | 0.0084 | 88.8 | 86.0 | 92.7 | 94.2 | 82.6 |

Optimal cutoffs of plasma pTau-217 and pTau-217/Aβ42 in differentiating CSF A/T groups were calculated considering the maximum sum of sensitivity and specificity as the metric. Abbreviations: NPV = negative predictive values, PPV = positive predictive value.

### 3.4 Plasma biomarkers across clinical groups

Figure 4 displays boxplots representing plasma levels of pTau-217, pTau-181, Aβ42/Aβ40, and pTau-217/Aβ42 ratios across diagnostic groups. AD-DEM patients exhibited significantly higher values of pTau-217, pTau-181, and pTau-217/Aβ42 compared to all other diagnostic groups, including AD-MCI subjects. Similarly, AD-MCI subjects displayed higher values compared to NonAD-MCI and NonAD-DEM, except for pTau-181, which showed comparable levels between AD-MCI and NonAD-DEM. Additionally, Aβ42/Aβ40 values were significantly lower in AD-DEM subjects compared to both NonAD-DEM and NonAD-MCI patients.

**Figure 4.**
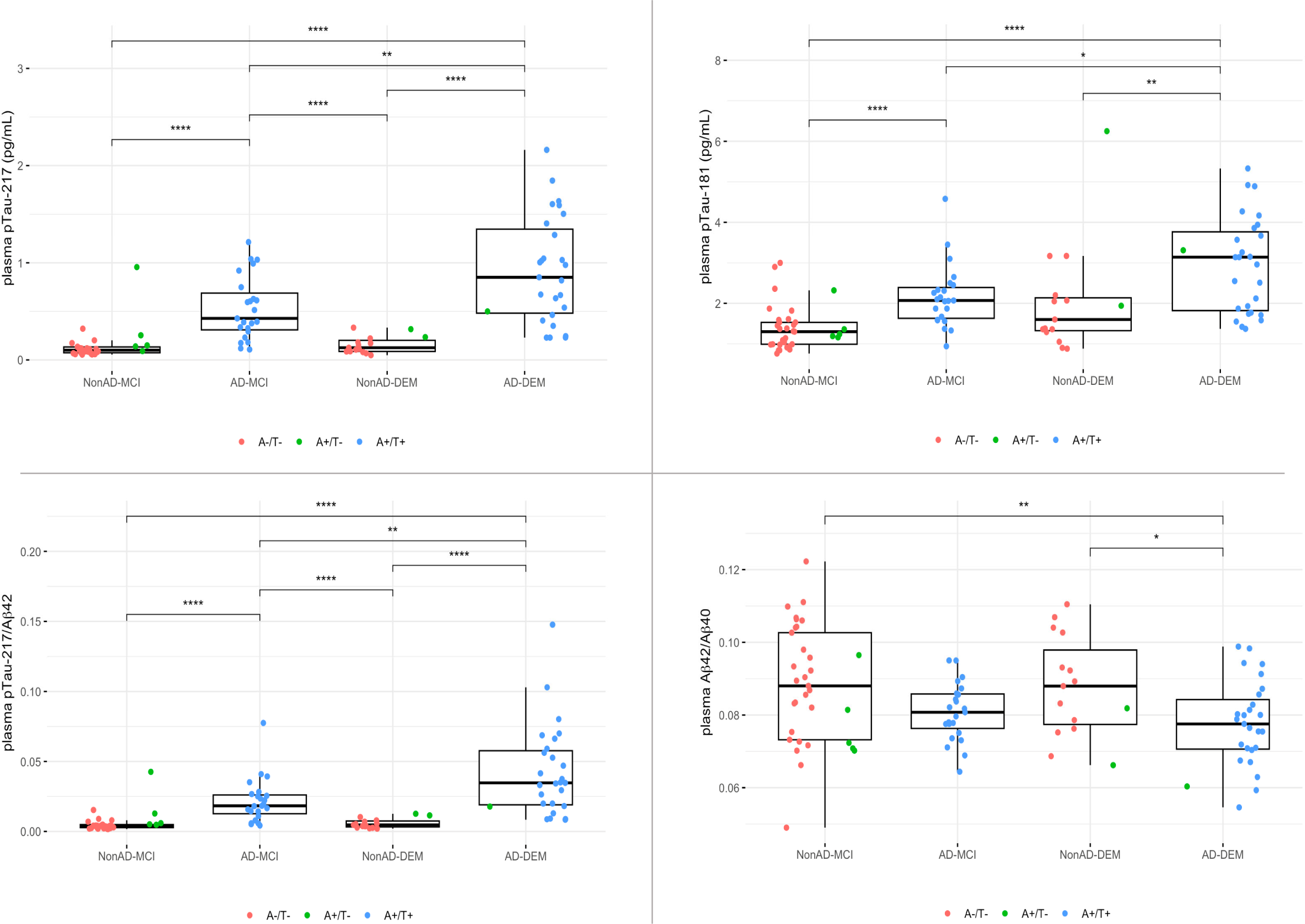
Plasma concentrations of Alzheimer’s disease (AD) biomarkers (pTau-217, pTau-181, pTau-217/Aβ42 ratio, Aβ42/Aβ40 ratio) across clinical diagnostic groups. The diagnostic groups include patients with Alzheimer’s dementia (AD-DEM), patients with mild cognitive impairment due to AD (AD-MCI), patients with non-AD related dementia (NonAD-DEM), and patients with non-AD related MCI (NonAD-MCI). The boxplots illustrate the data, with boxes representing the interquartile range, median concentrations depicted by horizontal lines within the boxes, and whiskers extending to the first/third quartile +/- 1.5 times the interquartile range. Pairwise comparisons were conducted on rank-transformed values using logistic regression, adjusting for age, sex, and kidney disease, with p-values Bonferroni-corrected for multiple comparisons.

ROC analysis (Figure 5) confirmed these differences in plasma biomarkers, with both pTau-217/Aβ42 and pTau-217 exhibiting excellent AUC values in reflecting Alzheimer’s neuropathology in both MCI and dementia clinical groups (AUCs: 0.93-0.97). Remarkably, the AUCs of pTau-217 and pTau-217/Aβ42 were significantly higher than those of pTau-181 (AUCs: 0.75-0.83) and Aβ42/Aβ40 (AUCs: 0.65-0.70) in both ‘AD-MCI vs NonAD-MCI’ and ‘AD-DEM vs NonAD-DEM’ comparisons. Moreover, pTau-181 AUC was significantly higher than Aβ42/Aβ40 AUC solely in the first comparison. Conversely, when attempting to reflect clinical stages within the AD continuum, AUC values were lower and comparable for all explored plasma biomarkers (AUCs: 0.60-0.73).

**Figure 5.**
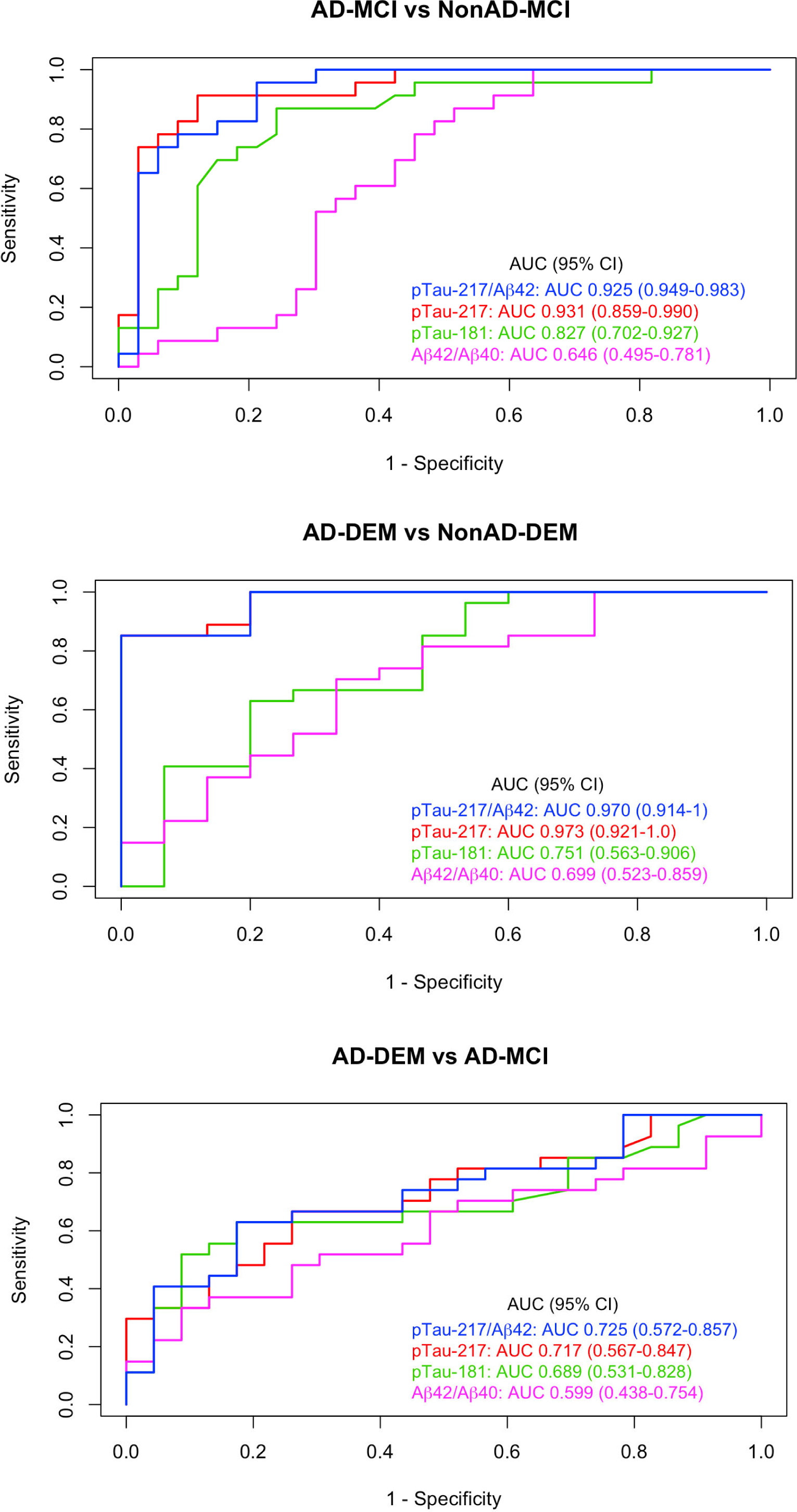
Receiver operating characteristic (ROC) curves assessing the discriminative ability of plasma biomarkers (pTau-217, pTau-181, pTau-217/Aβ42 ratio, Aβ42/40 ratio) in distinguishing among clinical diagnostic groups. Areas under the ROC curves (AUC) were calculated for each comparison, with their 95% confidence intervals generated by 2.000 bootstrap replicates. Plasma pTau-217 and pTau-217/Aβ42 ratio demonstrated excellent and superior diagnostic performance compared to pTau-181 and Aβ42/Aβ40 in discriminating patients with mild cognitive impairment AD-related (AD-MCI) and not related to AD (NonAD-MCI) and in distinguishing patients with dementia AD-related (AD-DEM) and not related to AD (NonAD-DEM).

## 4. Discussion

The present study builds on recent evidence highlighting the robust analytical and diagnostic capabilities of plasma pTau-217 measured using the fully automated Lumipulse G600II system. We conducted a comprehensive assessment and comparison of the performances of plasma pTau-217, pTau-181, Aβ42, Aβ40, and significant ratios. Our findings demonstrate that pTau-217 offers superior accuracy in reflecting CSF A/T status compared to plasma pTau-181 and the Aβ42/Aβ40 ratio. Interestingly, the ratio pTau-217/Aβ42 did not significantly improve the performance of pTau-217.

We found strong correlations between plasma levels of pTau-217 and CSF biomarker levels, particularly CSF pTau-181, supporting its potential use as a substitute for CSF examination.^9, 12^ It emerged as an accurate biomarker of AD, effectively distinguishing A+/T+ and A+ status with AUC values ranging from 0.94 to 0.97 (Figure 3), outperforming pTau-181 and Aβ42/Aβ40 ratio. Plasma pTau-217 ability to reflect AD was consistent across prodromal and dementia stages, showing AUC values of 0.93 and 0.97 at MCI and dementia stages, respectively (Figure 4 and 5). AD-DEM displayed the highest levels of pTau-217, consistent with prior evidence.^8^ This observation reasonably suggests an escalating rate of excretion from the central nervous system into the bloodstream as AD neuropathology progresses.^8^

Remarkably, identified cutoffs for plasma pTau-217 showed excellent accuracy, sensitivity and specificity, with a low misclassification rate (6-7%) for CSF A+/T+ status (Table 2). This suggests that the need for CSF analysis or amyloid PET scans could potentially be restricted to a subgroup of subjects with borderline levels.

We evaluated the diagnostic capabilities of the pTau-217/Aβ42 ratio to assess its potential additive value to pTau-217 alone. However, our analysis did not reveal any significant enhancement. This evidence reinforces the idea that plasma pTau-217 could be utilized as a standalone biomarker in clinical practice.^10^

Recent Italian studies have demonstrated comparable technical validity in Lumipulse plasma pTau-181 and Aβ42/Aβ40 ratio assays, consistent with our findings and with assays employing single molecule array (SIMOA) technology.^3, 6, 8^ It is worth noting that the slightly lower diagnostic accuracies of pTau-181 and Aβ42/Aβ40 ratio observed in our study may be partially attributed to our decision not to remove outliers, maintaining adherence to a real-life setting.

We examined possible confounders of plasma biomarkers levels in the entire sample and by subgrouping patients according to CSF A status. KD is known to exert substantial effects on plasma biomarkers performances, potentially mitigated by the use of ratios;^3, 8, 13^ consistently, our findings highlight a moderate association of KD with increased values of Aβ40 in all patients groups, as well as with increased levels of pTau-217 in CSF A- subjects, whereas correlations with biomarkers ratios (i.e., Aβ42/Aβ40, pTau-217/Aβ42) were weaker (Figure 1). Age exerted nonsignificant to weak effects on pTau-217, pTau-181 and Aβ42/Aβ40 ratio within the whole group and the A+ group. Within the A- groups, on the contrary, we found a moderate positive correlation of age with pTau-217 and with Aβ40; looking at the stricter association of GFR with age in the A- patients than in other groups (Figure 1), though, it might be hypothesized that KD may in larger part explain our findings.

Despite its strengths, our study has limitations. The sample size, while considerable, is lower than some other studies. However, the significance of our findings, coupled with consistency with recent studies, underscores the reliability of fully automated Lumipulse plasma biomarker assays across centers. Furthermore, we did not have data on blood-brain barrier permeability, which could have added further insights into plasma biomarkers dynamics.

Longitudinal studies are warranted to establish the prognostic significance of plasma pTau-217 in predicting clinical progression and its value as biomarker of therapeutic response in the emerging era of disease-modifying therapies for AD.^14^ Additionally, investigations into the underlying mechanisms regulating plasma biomarker levels and their correlation with neuroimaging and cognitive outcomes are warranted to further elucidate their role in AD pathophysiology.

By demonstrating the excellent and superior discriminative performance of plasma pTau-217 in reflecting CSF A+ and A+/T+ status in a real-life setting, our study addresses the ongoing need for reliable automated assays for AD diagnosis in clinical practice. These findings strongly support the potential of pTau-217 as a single, reliable non-invasive biomarker for the diagnosis of AD in real-world settings.

## Data Availability

All data produced in the present study are available upon reasonable request to the authors

## Acknowledgments

We express our gratitude to Fujirebio Italia for their invaluable support in providing the research kits free of charge for our scientific investigation.

## Abbreviations

AD: Alzheimer’s disease
AD-DEM: patients with Alzheimer’s dementia
AD- MCI: patients with mild cognitive impairment due to AD
CSF: cerebrospinal fluid
GFR: glomerular filtration rate
KD: kidney disease
NonAD-DEM: patients with dementia non-AD related
NonAD-MCI: patients with MCI non-AD related

## References

1. Jack CR, Jr., Bennett DA, Blennow K, et al. NIA-AA Research Framework: Toward a biological definition of Alzheimer’s disease. Alzheimers Dement 2018;14:535–562.

2. Ashton NJ, Brum WS, Di Molfetta G, et al. Diagnostic Accuracy of a Plasma Phosphorylated Tau 217 Immunoassay for Alzheimer Disease Pathology. JAMA Neurol 2024;81:255–263.

3. Bellomo G, Bayoumy S, Megaro A, et al. Fully automated measurement of plasma Abeta42/40 and p-tau181: Analytical robustness and concordance with cerebrospinal fluid profile along the Alzheimer’s disease continuum in two independent cohorts. Alzheimers Dement 2024.

4. Jonaitis EM, Janelidze S, Cody KA, et al. Plasma phosphorylated tau 217 in preclinical Alzheimer’s disease. Brain Commun 2023;5:fcad057.

5. Janelidze S, Bali D, Ashton NJ, et al. Head-to-head comparison of 10 plasma phospho-tau assays in prodromal Alzheimer’s disease. Brain 2023;146:1592–1601.

6. Quaresima VP, A.; Trasciatti, C.; Tolassi, C.; Parigi, M.; Bertoli, D.; Mordenti, C.; Galli, A.; Rizzardi, A.; Caratozzolo, S.; Benussi, A.; Ashton, N.J.; Blennow, K.; Zetterberg, H.; Giliani, S.C.; Brugnoni, D.; Padovani, A. Plasma P-Tau181 and Amyloid Markers in Alzheimer’s Disease: A Method Comparison between Simoa and Lumipulse 2024.

7. Mendes AJ, Ribaldi F, Lathuiliere A, et al. Head-to-head study of diagnostic accuracy of plasma and cerebrospinal fluid p-tau217 versus p-tau181 and p-tau231 in a memory clinic cohort. J Neurol 2024;271:2053–2066.

8. Arranz J, Zhu N, Rubio-Guerra S, et al. Diagnostic performance of plasma pTau 217, pTau 181, Abeta 1-42 and Abeta 1-40 in the LUMIPULSE automated platform for the detection of Alzheimer disease. Res Sq 2023.

9. Pilotto AQ, V.; Trasciatti, C.; Tolassi, C.; Bertoli, D.; Mordenti, C.; Galli, A.; Rizzardi, A.; Caratozzolo, S.; Zancanaro, A.; Contador, J.; Hansson, O.; Palmqvist, S.; De Santis, G.; Zetterberg, H.; Blennow, K.; Brugnoni, D.; Suárez-Calvet, M.; Ashton, N.J.; Padovani A. Plasma p-tau217 in Alzheimer’s disease: Lumipulse and ALZpath SIMOA head-to-head comparison. 2024.

10. Alzheimer’s Association Workgroup. Revised Criteria for Diagnosis and Staging of Alzheimer’s Disease [online]. Available at: https://aaic.alz.org/diagnostic-criteria.asp.

11. Mazzeo S, Santangelo R, Bernasconi MP, et al. Combining Cerebrospinal Fluid Biomarkers and Neuropsychological Assessment: A Simple and Cost-Effective Algorithm to Predict the Progression from Mild Cognitive Impairment to Alzheimer’s Disease Dementia. J Alzheimers Dis 2016;54:1495–1508.

12. Hansson O, Edelmayer RM, Boxer AL, et al. The Alzheimer’s Association appropriate use recommendations for blood biomarkers in Alzheimer’s disease. Alzheimers Dement 2022;18:2669–2686.

13. Janelidze S, Barthelemy NR, He Y, Bateman RJ, Hansson O. Mitigating the Associations of Kidney Dysfunction With Blood Biomarkers of Alzheimer Disease by Using Phosphorylated Tau to Total Tau Ratios. JAMA Neurol 2023;80:516–522.

14. Teunissen CE, Verberk IMW, Thijssen EH, et al. Blood-based biomarkers for Alzheimer’s disease: towards clinical implementation. Lancet Neurol 2022;21:66–77.

